# Immunogenic profile of SARS-CoV-2 spike in individuals recovered from COVID-19

**DOI:** 10.1101/2020.05.17.20104869

**Authors:** Jennifer A Juno, Hyon-Xhi Tan, Wen Shi Lee, Arnold Reynaldi, Hannah G Kelly, Kathleen Wragg, Robyn Esterbauer, Helen E Kent, C Jane Batten, Francesca L Mordant, Nicholas A Gherardin, Phillip Pymm, Melanie H Dietrich, Nichollas E Scott, Wai-Hong Tham, Dale I Godfrey, Kanta Subbarao, Miles P Davenport, Stephen J Kent, Adam K Wheatley

## Abstract

The rapid global spread of SARS-CoV-2 and resultant mortality and social disruption have highlighted the need to better understand coronavirus immunity to expedite vaccine development efforts. Multiple candidate vaccines, designed to elicit protective neutralising antibodies targeting the viral spike glycoprotein, are rapidly advancing to clinical trial. However, the immunogenic properties of the spike protein in humans are unresolved. To address this, we undertook an in-depth characterisation of humoral and cellular immunity against SARS-CoV-2 spike in humans following mild to moderate SARS-CoV-2 infection. We find serological antibody responses against spike are routinely elicited by infection and correlate with plasma neutralising activity and capacity to block ACE2/RBD interaction. Expanded populations of spike-specific memory B cells and circulating T follicular helper cells (cTFH) were detected within convalescent donors, while responses to the receptor binding domain (RBD) constitute a minor fraction. Using regression analysis, we find high plasma neutralisation activity was associated with increased spike-specific antibody, but notably also with the relative distribution of spike-specific cTFH subsets. Thus both qualitative and quantitative features of B and T cell immunity to spike constitute informative biomarkers of the protective potential of novel SARS-CoV-2 vaccines.

## Introduction

The rapid global spread of SARS-CoV-2 has highlighted the intrinsic vulnerability of humans to emerging zoonotic infections and spurred frantic efforts to expedite vaccine and antiviral drug development, manufacture and deployment. In contrast to historical pandemics, such as the 1918 “Spanish” H1N1 influenza, modern recombinant technology enables a rapid scientific response, with multiple vaccines under development, almost exclusively aimed at eliciting antibodies to the viral “spike” protein.

The spike (S) protein of beta-coronaviruses is expressed as a single protein, with proteolytic cleavage yielding S1 and S2 subunits ^1^. S localises on the virion surface and mediates both recognition of cellular receptors and membrane fusion. In the case of SARS-CoV-2, a receptor binding domain (RBD) within S1 directly interacts with high affinity with the peptidase domain of angiotensin-converting enzyme 2 (ACE2) ^2–4^ The S2 subunit of S mediates membrane fusion. The S/ACE2 interaction mediates viral entry and provides an attractive target for vaccine-elicited humoral immunity ^5^, with antibodies potentially capable of either (i) directly blocking binding of ACE2 by S, (ii) blocking conformational changes in S critical for membrane fusion, (iii) eliminating infected cells through antibody effector mechanisms such as antibody-dependent cellular cytotoxicity (ADCC), or (iv) driving accelerated clearance of free virus.

The dominant targets for human antibody against the SARS-CoV-2 S are unclear. Some human mAbs originally characterised against SARS-CoV S cross-react with SARS-Cov-2. For example CR3022 which binds a cryptic epitope on the RBD ^6,7^, while S309, derived from the memory B cells of a SARS-CoV recovered subject, blocks ACE2 engagement by SARS-CoV-2 S ^8^. A recent report of monoclonal antibodies recovered from SARS-CoV-2 convalescent donors revealed multiple non-overlapping epitopes on the RBD, with different capacities for mediating neutralisation^9^. Few neutralising epitopes localised outside the RBD have been characterised to date, with preliminary reports of neutralising epitopes within the N-terminal domain of S ^10^. Nevertheless, studies of other zoonotic human coronaviruses MERS-CoV and SARS-CoV have identified epitopes in both the N-terminal domain of S1 or within S2 can neutralise virus infection by blocking critical viral functions such as membrane fusion^11–14^.

Given the capacity to prevent viral infection, eliciting antibodies to S by immunisation is of intense interest and multiple human vaccines targeting S are in various stages of clinical development (reviewed in ^15,16^). However, although an inactivated SARS-CoV-2 vaccine showed promise in animal models ^17^, the immunogenic properties of SARS-CoV, MERS-CoV or SARS-CoV-2 S-based immunogens in humans are poorly resolved. Here we undertook an in-depth characterisation of humoral and cellular immunity against spike in humans who recovered from mild to moderate SARS-CoV-2 infection. We find antibody responses to both S and the RBD are consistently elicited following SARS-CoV-2 infection, the magnitude of which correlates with both plasma neutralising activity and inhibition of RBD/ACE2 binding. S-specific B cells comprise a significant proportion of the circulating memory B cell pool following infection, with RBD-specific B cells constituting a minor subpopulation in most subjects. Assessment of the circulating T follicular helper (cTFH) population reveals that S-specific cTFH cells are also readily detected in convalescent subjects, while T cell responses toward the RBD are significantly lower in frequency. Finally, we find the development of comparatively high plasma neutralisation activity is associated not only with the magnitude of anti-S immune responses, but also with the phenotype of circulating TFH populations, suggesting these features may serve as attractive biomarkers for candidate S-based vaccines entering the clinic.

## Results

### Serological responses to spike antigens following SARS-CoV-2 infection

We recruited a cross-sectional cohort (N=41) of Australian adults recovered from mild-moderate SARS-CoV-2 infection and isolated plasma and PBMC samples at a median of 32 (IQR: 28-35) days post-positive PCR test. The cohort had a median age of 59 (IQR: 54-65) and was 43% female (17 of 41). Subjects reported mild to moderate upper and lower respiratory tract symptoms with only 5 (12%) requiring hospitalisation, and none requiring mechanical ventilation (Table S1). A control cohort of 27 healthy adults was recruited prior to widespread infection in Australia (Table S2). As we had an interest in the degree to which baseline cross-reactive coronavirus immunity affected SARS-CoV-2 responses, we pre-screened the 27 uninfected subjects for serological reactivity against the beta coronavirus HCoV-HKU1 (HKU1) (Figure S1), selecting individuals with the 5 highest and 5 lowest plasma titres as controls for the study.

Serological profiles are presented stratified across the cohort based on neutralisation activity for each subject. Antibodies binding the SARS-CoV-2 spike (Figure 1A) or the RBD (Figure 1B) were consistently observed in all 41 infected individuals by ELISA, with minimal reactivity in the controls. Titres of S- and RBD-specific antibody were highly correlated (Figure S2). Consistent with previous reports ^18^, low-level antibody responses cross-recognising the SARS-CoV RBD were observed in most of our SARS-CoV-2 infected cohort (Figure 1C). Antibody responses to the human coronavirus strain HKU were prevalent at moderate to high levels across the cohort, in line with previous reports of widespread seropositivity to S proteins of human coronaviruses in adults ^19,20^ (Figure 1D). The capacity of immune plasma to block interaction between recombinant ACE2 and RBD was assessed by ELISA, with modest levels of inhibition detected in most subjects, and selected subjects exhibiting potent inhibitory activity (Figure 1E). Virus neutralising activity in the plasma was similarly assessed using a microneutralisation assay with live SARS-CoV-2 infection of Vero cells as previously described for SARS-CoV and MERS-CoV^21,22^. Neutralising antibody titres ranged from 40 to 508 (IQR:113-254) (Figure 1F). In summary, antibody responses against both S and the RBD are consistently elicited in SARS-CoV-2 infected individuals, the endpoints titres of which correlate significantly with neutralising activity (r=0.55 and r=0.61 respectively) and ACE2 binding inhibition (r=0.72 and r=0.72 respectively) in the plasma (Figure S2).

**Figure 1.**
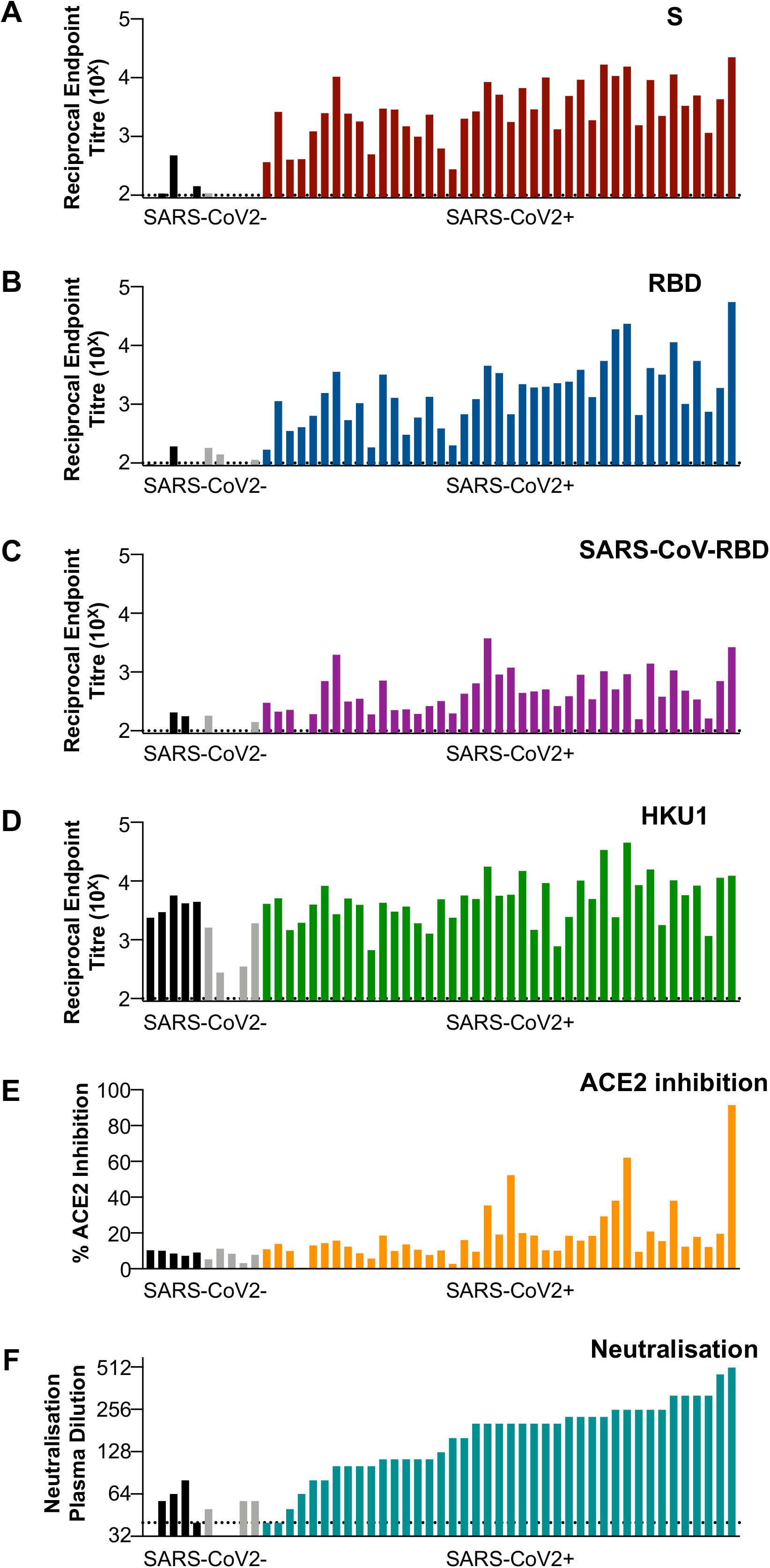
Serological responses to COVID19. Plasma samples from subjects recovered from SARS-CoV-2 infection (N=41) and healthy controls (N=10, black/grey bars) were screened by ELISA for reactivity against S recombinant proteins from (**A**) SARS-CoV-2 S, (**B**) SARS-CoV-2 RBD, (**C**) SARS-CoV RBD and (**D**) HCoV-HKU1. (**E**) The capacity of plasma antibodies to inhibit the interaction of RBD with human ACE2 was tested in an ELISA format at a plasma dilution of 1:25. (**F**) Neutralisation activity in the plasma was assessed using a microneutralisation assay. Data represent individual responses for each subject, and column order for all figures is conserved and stratified on the basis of plasma neutralisation activity. SARS-CoV-2 negative donors were stratified based on HCoV-HKU1 serological titres (black, high titres; grey, low titres).

### B cell responses to S antigens following SARS-CoV-2 infection

We next examined the frequency and specificity of class-switched B cells in convalescent subjects using SARS-CoV-2 spike or RBD proteins as flow cytometric probes. Clear antigen-specific populations of CD19^+^IgD^−^ B cells (gating in Figure S3) binding spike, spike and RBD or RBD alone could be resolved in our cohort of recovered from SARS-CoV-2 subjects, with minimal background staining in uninfected controls (Figure 2A; Figure S4). Frequencies of spike^+^RBD^−^, spike^+^RBD^+^ and spike^−^RBD^+^ B cells as a proportion of the CD19^+^IgD^−^ population were a median 0.38% (IQR 0.24-0.52), 0.047% (IQR 0.023-0.084) and 0.033% (IQR 0.015-0.045), respectively (Figure 2B). The very low frequencies of spike^−^RBD^+^ B cells likely constitute a mix of background staining and B cells that recognise RBD epitopes occluded in recombinant S or intact virus. Immunoglobulin isotypes were determined for spike^+^RBD^−^ and spike^+^RBD^+^ populations using IgM and IgG surface staining, with IgM^−^IgG^−^ class-switched B cells previously established to be almost exclusively IgA^+^ ^23^. In our cohort sampled a median of 36 days after symptom onset, the majority of spike^+^RBD^−^ class-switched B cells were IgG^+^ (median 57.5%; IQR 46.8-64.8), with smaller proportions displaying IgM^+^ (20.9%; IQR 17.4-29.1) and IgA^+^ (IgM^−^IgG^−^) (17.4%; IQR 13.2-25.9) (Figure 2C). Isotype distribution of spike^+^RBD^+^ B cells was more variable due to low event counts, with median frequencies of 45.5% (IQR 27.8-70.7) IgG, 13.6% (IQR 0-30.3) IgM and 20.0% IgA (IQR 4.73-41.9). The activation phenotype of antigen-specific B cells was assessed using CD21 and CD27 surface staining^24^(Figure S5). Most spike^+^RBD^−^ (median 58.5%; IQR 52.2-66.6) or spike^+^RBD^+^ (68.7%; IQR 54.4-80) class-switched B cells displayed a resting memory phenotype (CD21^+^CD27^+^), also consistent with the median duration of infection. However, a significant proportion of activated memory B cells (CD21 ^−^CD27^+^) was still evident for both spike^+^RBD^−^ (18.9%; IQR 13.2-25.7) or spike^+^RBD^+^ B cell populations (13.2%; IQR 5.88-20), with only low proportions of CD21^−^CD27^−^ and CD21^+^CD27^−^ phenotypes observed. Overall, SARS-CoV-2 infection efficiently elicits both S- and RBD-specific B cells in most subjects after recovery, which constitute a significant proportion of the memory B cell pool, which are mostly IgG^+^ and of a resting memory phenotype.

**Figure 2.**
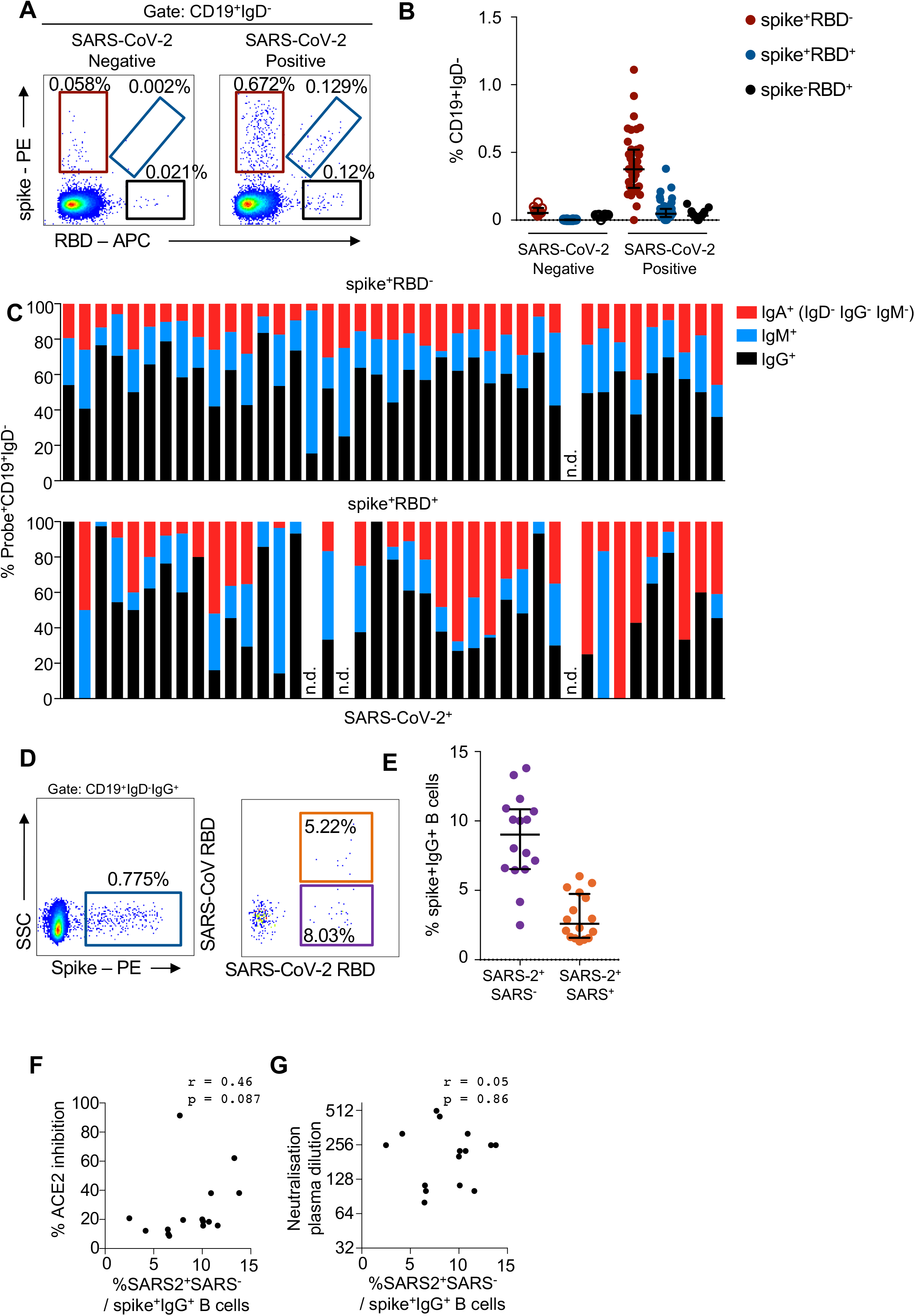
Frequency and phenotype of SARS-CoV-2-specific B cells following infection. (**A**) Co-staining class-switched B cells (CD19^+^IgD-) with SARS-CoV-2 spike and RBD probes allows resolution of antigen-specific cells in subjects previously infected with SARS-CoV-2 relative to uninfected controls. (**B**) Frequencies of spike^+^RBD^−^, spike^+^RBD^+^ and spike^+^RBD^−^ B cells as a proportion of CD19^+^IgD^−^ B cells in PBMC from subjects previously infected with SARS-CoV-2 (N=41) and uninfected controls (N=10). (**C**) Isotype distribution of spike^+^RBD^−^ and spike^+^RBD^+^ CD19^+^IgD^−^ B cells; n.d – not detected due to absent probe^+^ cells. (**D**) Co-staining spike^+^CD19^+^IgD^−^ B cells with SARS-CoV-2 and SARS-CoV RBD probes allows discrimination of cross-reactive specificities versus those unique to SARS-CoV-2. **(E)** Frequency of SARS-CoV-2 RBD^+^/SARS-CoV RBD^−^ cells as a proportion of spike^+^CD19^+^IgD-IgG^+^ B cells and their correlation (Spearman) to (**F**) ACE2/RBD binding inhibition and (**G**) neutralising activity in plasma.

The RBD of SARS-CoV and SARS-CoV-2 share significant homology, but with marked diversity within the ACE2 binding motif despite shared recognition of this cellular receptor ^2,25^. We examined whether differential staining with SARS-CoV and SARS-CoV-2 probes would allow more precise identification of B cells recognising the unique ACE2 binding site of SARS-CoV-2, to understand why some individuals had notable RBD-specific antibody titres but with limited neutralisation activity or RBD/ACE2 binding inhibition. PBMCs from a subset of COVID^+^ subjects (N=15) were stained with SARS-CoV-2 spike, SARS-CoV-2 RBD and SARS-CoV RBD probes as before (Figure 2D). Both SARS-CoV-2 RBD-specific and SARS-CoV/SARS-CoV-2 cross-reactive IgG^+^ B cells could be resolved in most subjects (Figure S6), with cells binding only SARS-CoV-2 RBD making up the major fraction of the spike^+^RBD^+^ response (Figure 2E). However, while increased proportions of SARS-CoV-2 RBD-specific B cells within the spike^+^ response does broadly track with plasma blockade of RBD/ACE2 binding (Figure 2F; r=0.46), we find no correlation with neutralisation activity (Figure 2G: r=0.05).

### CD4 T cell responses to S antigens following SARS-CoV-2 infection

TFH provide critical cognate help to antigen-specific B cells to initiate and maintain humoral immune responses ^26^. Circulating TFH cells (cTFH; CD3^+^CD4^+^CD45RA^−^ CXCR5^+^, gating in Figure S7) in the blood provide a surrogate indication of TFH activity in lymphoid tissues ^27,28^. Total unstimulated cTFH frequencies were similar across SARS-CoV-2 convalescent and uninfected donors (Figure S8). Antigen specificity of the cTFH population was determined using an activation induced marker (AIM) assay ^29^ in response to stimulation with SARS-CoV-2 spike or RBD proteins (Figure 3A). Overall, recovered subjects exhibited robust cTFH responses to the SARS-CoV-2 spike protein, with a median of 0.92% spike-specific cTFH cells (IQR 0.42 – 1.52; Figure 3B). In contrast to the full spike, RBD-specific cTFH responses were significantly lower (p<0.0001), with a median of only 0.12% of cTFH cells exhibiting RBD specificity (IQR 0.04 – 0.38). Consistent with the high frequency of HKU1 seropositivity among the convalescent cohort (Figure 1D), cTFH responses to HKU1 spike were detected among 97.5% of donors (median 0.52% of cTFH cells, IQR 0.32 – 0.99). The frequency of HKU1-specific cTFH was generally higher among the convalescent cohort than the uninfected controls (Figure 3B), suggesting a boosting of cross-reactive T cell responses following SARS-CoV-2 infection.

**Figure 3.**
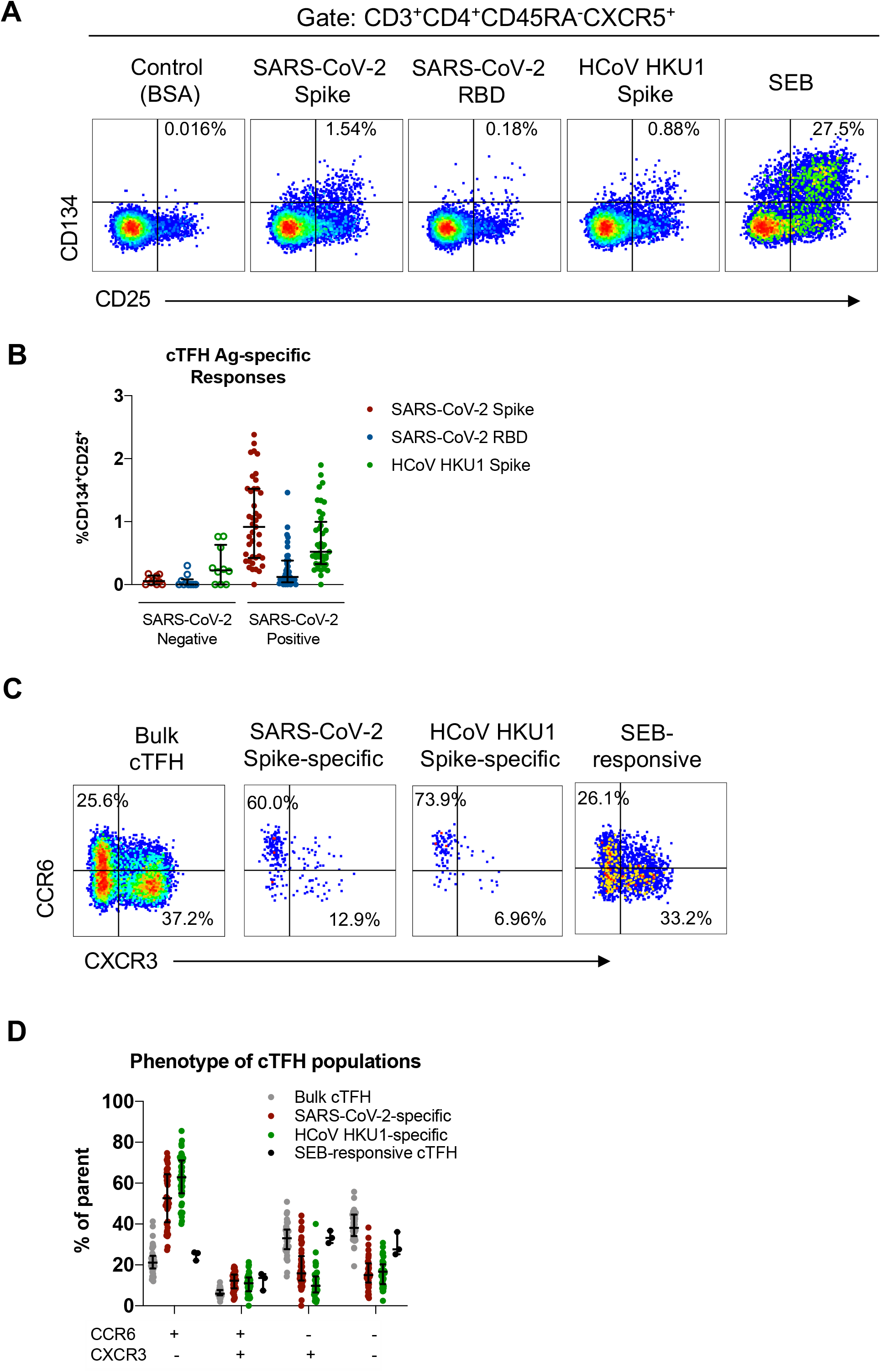
Specificity of cTFH responses to coronavirus spike proteins. (**A**) Representative staining of CD25 and CD134 co-expression on cTFH (CD3^+^CD4^+^CD45RA^−^CXCR5^+^) cells following stimulation with 5μg/mL BSA (negative control), SARS-CoV-2 Spike, SARS-CoV-2 RBD or HCoV HKU1 protein, or SEB (positive control). (**B**) Antigen-specific cTFH (n=10 SARS-CoV-2 negative, n=41 SARS-CoV-2 positive donors) frequencies were calculated as the proportion of CD25^+^CD134^+^ cTFH cells in each stimulation condition after background subtraction using the negative control. (**C**) Representative expression of CCR6 and CXCR3 on bulk cTFH, SARS-CoV-2 Spike-specific, HCoV HKU1 Spike-specific or SEB-responsive (CD25^+^OX^−^40^+^) cTFH. (D) Quantification of CCR6^+^CXCR3^−^, CCR6^+^CXCR3^+^, CCR6^−^CXCR3^+^ or CCR6^−^CXCR3^−^ cTFH populations among SARS-CoV-2 positive donors (n=41).

cTFH populations have classically been divided on the basis of surface expression of chemokine receptors CCR6 and CXCR3 ^28,30,31^. However the functional capacities and relevance of putative cTFH1 (CCR6^−^CXCR3^+^), cTFH2 (CCR6^−^CXCR3^−^) and TH17-like (CCR6^+^CXCR3^−^) cTFH subsets are still unclear and may be pathogen-dependent or vary with anatomical location ^27,28,31^. In contrast to the total unstimulated cTFH population, SARS-CoV-2 spike and HKU-specific cTFH were enriched for CCR6^+^CXCR3^−^ cells (median 52.6% of SARS-CoV-2 spike-specific, 62.9% of HKU1 spike-specific or 21.1% of bulk cTFH, p<0.0001, Figure 3C/3D). Comparison to SEB-stimulated cells from a subset of donors confirmed that *in vitro* TCR stimulation does not preferentially activate or upregulate expression of CCR6 among the cTFH population (Figure 3D).

Analysis of spike-specific non-cTFH CD4 memory (CD3^+^CD4^+^CD45RA^−^CXCR5^−^) cells revealed similar patterns of antigen reactivity to the cTFH compartment; namely, strong recognition of SARS-CoV-2 and HKU1 spike proteins (median 0.53% and 0.54% of CD4 memory cells, respectively) and lower frequencies of RBD-specific T cells (median 0.24% of CD4 memory cells) (Figure S8).

### Predictors of plasma neutralisation activity

The development of serological neutralisation activity will be a critical endpoint for upcoming SARS-CoV-2 vaccine trials. A co-correlation matrix of subject characteristics and immunological parameters was generated (Figure 4A). This analysis highlighted broad co-correlation of many immune parameters related to S immunogenicity, namely antibody titres and the circulating frequencies of S-specific B and T cell populations. Principal component analysis (PCA) on immunological variables revealed clustering of the cohort into subjects with stronger and weaker plasma neutralisation activity (Figure 4B). Using a multiple regression approach, we identified titres of S-specific antibody and the proportion of S-specific cTFH with a TH17-like phenotype (CCR6^+^CXCR3^−^) as the two most significant predictive factors related to neutralisation activity (Figure 4C).

**Figure 4.**
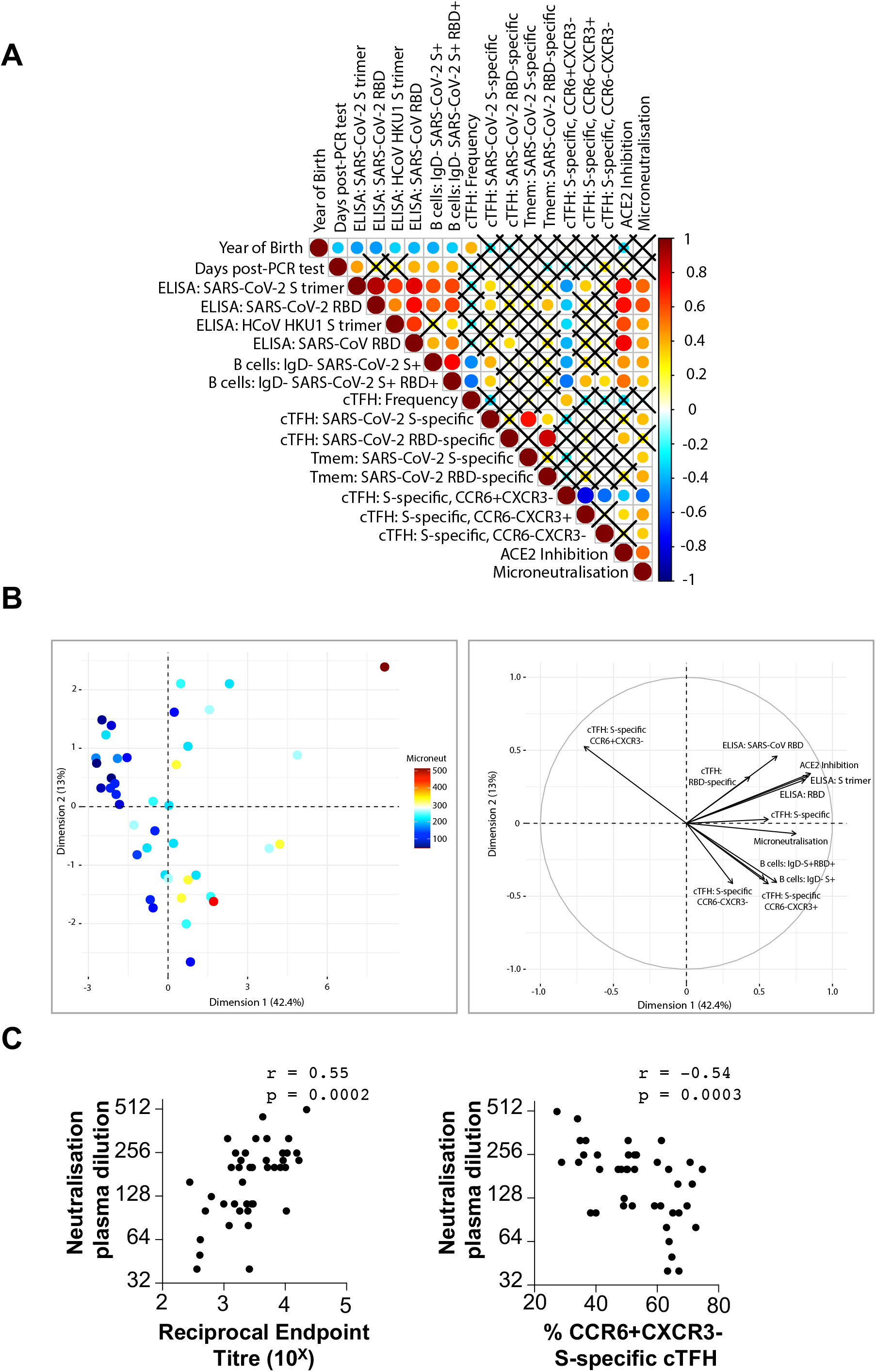
Predictors of plasma neutralisation activity. (**A**) Co-correlation matrix of subject characteristics, infection and immunological dynamics in subjects recovered from SARS-CoV-2 infection. (**B**) Principle component analysis of immune parameters showing individuals coloured in proportion to plasma neutralization titres. (**C**) Spearman correlation of the plasma neutralisation activity with S-specific antibody titres and the proportion of CCR6^+^CXCR3^−^ phenotype with the S-specific cTFH population.

## Discussion

Efficient elicitation of potent antibodies capable of neutralising viral entry is likely to be a critical feature of effective vaccines against SARS-CoV-2. In the current study, we observed that neutralisation activity in the plasma of convalescent subjects ranged from potent to negligible, despite near universal detection of antibodies binding S and/or RBD, suggesting that qualitative aspects of the humoral immune response may be a critical consideration for vaccine development. Direct assessment of key immunological events within the respiratory tract and draining lymphoid tissues is challenging in humans, however assessing B and T cell immunity in more readily sampled blood can be informative.

Spike-specific class-switched B cells were expanded in nearly all infected subjects, with a predominantly IgG^+^ and resting memory phenotype consistent with the sampling time several weeks after the resolution of infection. B cells binding the RBD, which contains the ACE2 interaction site, were markedly less frequent than S-specific B cells, and not detected at all in many subjects. Combinatorial B cell staining with both SARS-CoV and SARS-CoV-2 probes enabled focused assessment of the uniquely variant epitope on the SARS-CoV-2 RBD that facilitates high affinity recognition of ACE2. A minority of SARS-CoV-2 RBD-specific B cells also recognise the SARS-CoV RBD, a finding consistent with the relative infrequency of SARS-CoV or MERS-CoV cross-reactive antibodies recovered from convalescent patients to date^9,32^. We find the frequency of IgD^−^IgG^+^ B cells that bound S and SARS-CoV-2 RBD, but not cells binding SARS-CoV RBD, tracked with serological RBD/ACE2 binding inhibition but not with overall neutralising activity. Overall, our data suggest that in some subjects, precise antibody recognition and blockade of the RBD ACE2-binding site is the principal pathway to generating neutralising antibody. However, the disconnect seen in many subjects between plasma neutralising titres and RBD-specific antibody, B and T cell responses, strongly suggests sufficient non-RBD epitope targets exist to constitute an alternative pathway to comparable virus neutralisation outcomes.

TFH cells provide critical help to antigen-specific B cells in lymph nodes through contact-dependent (ICOS, CD154) and independent (IL-21) mechanisms ^26^. cTFH (CD45RA^−^CXCR5^+^ CD4 T cells) in the circulation have been suggested to serve as a surrogate measure of TFH activity ^28^. We found the frequency of SARS-CoV2 S-specific cTFH elicited following infection correlated with both S-specific antibody responses and S-specific B cell frequencies. In contrast, only limited cTFH responses to SARS-CoV-2 RBD were observed, which may reflect limited CD4 T cell epitopes given the small size of RBD. This has implications for RBD-based vaccine strategies, with inefficient recruitment of T cell help potentially limiting effective humoral responses, as previously observed with the stem domain of influenza hemagglutinin^33^.

Antigen-specific cTFH can be phenotypically characterised by surface expression of CXCR3 and CCR6. Studies suggest that CCR6^+^ cTFH cells produce more IL-21, provide superior help to naïve B cells and more transcriptionally resemble GC TFH ^27^, while CXCR3^+^ cTFH can preferentially provide help to memory B cells ^28,31^. We find SARS-CoV-2 S-specific cTFH exhibit a clear phenotypic bias toward a CCR6^+^CXCR3^−^ phenotype (cTFH17), consistent with responses to other neo-antigens such as Ebola glycoprotein vaccines ^34^. However despite this predominance, the relative proportion of S-specific cTFH17 (CCR6^+^CXCR3^−^) was negatively correlated with virus neutralisation activity. In contrast, increased frequencies of both cTFH1 (CCR6^−^CXCR3^+^) and cTFH2 (CCR6^−^CXCR3^−^) were observed in subjects with the highest plasma neutralising activity. Expansion of cTFH1 is well characterized following seasonal influenza immunisation, where peak frequencies in the blood correlate with both plasmablast expansion and subsequent serum neutralising antibody titres ^23,30,35^. Similarly, bias toward CXCR3^+^ phenotypes is reported for antigen-specific cTFH in many chronic infections ^36,37^. The functional significance of CXCR3^+^ cTFH during SARS-CoV-2 infection is currently unclear, however may reflect differences in lymph node TFH activity or egress from the GC.

The impact of widespread pre-existing immunity to human coronaviruses (229E, NL63, HKU1, OC43) upon the responses to SARS-CoV-2 infection is an open question. Here we found serum antibody against HKU1 was widely prevalent, consistent with the high seroprevalence rates in adults reported previously ^19,20^. However, we see no evidence of HKU-specific immunity modulating binding or neutralising titres against SARS-CoV-2 antigens. Our data suggest CD4 T cell responses to HKU1 may be boosted following SARS-CoV-2 infection, possibly via recognition of conserved epitopes within the S2 domain ^38^. The predominantly CCR6^+^ phenotype of SARS-CoV-2 and HKU-1-specific cTFH may reflect a coronavirus-specific TFH response, but further epitope mapping is required to deconvolute the contribution of HKU1 memory responses or recently boosted SARS-CoV-2 cross-reactivity.

There is understandably considerable scientific interest in predicting the biogenesis of protective immunity against SARS-CoV-2, of which neutralising antibodies against S are likely to be consequential. Although the current study is limited by cohort size, we find that concomitant factors demarking robust humoral immunity, namely increased S-specific class-switched B cells, circulating TFH and antibody, all correlate with increased virus neutralisation activity. Interestingly, we find the surface phenotype of circulating TFH populations most clearly differentiated subjects with potent neutralising responses. We propose B cell and cTFH frequencies and phenotypes constitute informative biomarkers of immune function for assessment of upcoming clinical trials of novel vaccines targeting S.

## Materials and Methods

### Ethics Statement

The study protocols were approved by the University of Melbourne Human Research Ethics Committee (#2056689) and all associated procedures were carried out in accordance with the approved guidelines. All participants provided written informed consent in accordance with the Declaration of Helsinki.

### Subject recruitment and sample collection

Subjects who had recovered from COVID19 and healthy controls were recruited through contacts with the investigators and invited to provide a blood sample. Subject characteristics of SARS-CoV-2 convalescent subjects are collated in Table S1 and the healthy controls in Table S2. For all participants, whole blood was collected with sodium heparin anticoagulant. Plasma was collected and stored at −80°C, and PBMCs were isolated via Ficoll Paque separation, cryopreserved in 10% DMSO/FCS and stored in liquid nitrogen.

### Expression of coronavirus proteins and hACE2

A set of proteins was generated for serological and flow cytometric assays. The ectodomain of SARS-CoV-2 (isolate WHU1;residues 1 – 1208) or HCoV-HKU1 S protein (isolate N5;residues 1 – 1290) were synthesised with furin cleavage site removed and P986/987 stabilisation mutations^39^, a C-terminal T4 trimerisation domain, Avitag and His-tag, expressed in Expi293 cells and purified by Ni-NTA affinity and size-exclusion chromatography using a Superose 6 16/70 column (GE Healthcare) (Figure S9). SARS-CoV S was biotinylated using Bir-A (Avidity). The SARS-CoV-2 RBD^40^ with a C-terminal His-tag (residues 319-541; kindly provided by Florian Krammer) was similarly expressed and purified. SARS-CoV RBD (residues N321-P513) with a C-terminal Avitag and His-tag, was expressed in Expi293 cells and purified by Ni-NTA, biotinylated using Bir-A (Avidity) and purified by ize-exclusion chromatography using a S-75 Superdex (GE Healthcare). The human (residues 19-613) and mouse (residues 19-615) ACE2 ectodomain with C-terminal His-tag (kindly provided by Merlin Thomas) were expressed in Expi293 cells and purified using Ni-NTA and size-exclusion chromatography (Figure S10). Antigenicity of coronaviral proteins was assessed by binding to immune sera, anti-RBD mAbs CR3022 and 4B, or human and mouse ACE2 (Figure S11). The glycosylation profile of recombinant S proteins (Figure S11) was assessed using mass spectrometry as previously described^41^ by SP3 protein clean up^42^ and trypsin in-solution digestion. Purified peptides were desalted then separated using a two-column chromatography set up comprising a PepMap100 C18 20 mm × 75 μm trap and a PepMap C18 500 mm × 75 μm analytical column on Dionex Ultimate 3000 UPLC (ThermoFisher). Samples were concentrated onto the trap column at 5 μl/min with Buffer A (2% acetonitrile, 0.1% formic acid) for 6 min and infused into a Q-Exactive™ plus mass spectrometry (ThermoFisher) at 300 nl/min via the analytical column. 125 min gradients were used altering the buffer composition from 2% Buffer B (80% acetonitrile, 0.1% formic acid) to 28% B over 95 min, then from 28% B to 40% B over 10 min, then from 40% B to 100% B over 2 min, the composition was held at 100% B for 3 min, and then dropped to 3% B over 5 min and held at 3% B for another 10 min. The Q-Exactive™ plus Mass Spectrometer was operated in a data-dependent mode automatically switching between the acquisition of a single Orbitrap MS1 scan (70,000 resolution, AGC of 3 × 10^6^) followed by 15 data-dependent HCD MS2 events (35,000 resolution; stepped NCE 28, 30, 32; maximum injection time of 125 ms and AGC of 2 × 10^5^) with 30 s dynamic exclusion enabled.

The identification of glycoforms were accomplished using Byonic [Protein Metrics, version 3.5.3^43^. MS raw file were searched with a MS1 tolerance of ±5 ppm and a tolerance of ±20 ppm was allowed for HCD MS2 scans. Searches were performed using cysteine carbamidomethylation as a fixed modification, methionine oxidation as a variable modification in addition to allowing *N*-linked glycosylation on asparagine. The default Byonic *N*-linked glycan database, which is composed of 309 mammalian *N*-glycans compiled was used. The proteases specificity was set to full trypsin specificity and a maximum of two miss-cleavage events allowed. Data searched against the expected protein sequence. Search was filtered to a 1% protein FDR as set in the Byonic parameters with the final results filtered to remove glycopeptide assignments with Byonic score below 300 [double the cut off score suggested by Lee *et al^44^*] to remove low quality glycopeptide assignments. Data are available via ProteomeXchange with identifier PXD019163.

### ELISA

Antibody binding to coronavirus S or RBD proteins was tested by ELISA. 96-well Maxisorp plates (Thermo Fisher) were coated overnight at 4°C with 2_μg/mL recombinant S or RBD proteins. After blocking with 1% FCS in PBS, duplicate wells of serially diluted plasma were added and incubated for two hours at room temperature. Plates were washed prior to incubation with 1:20000 dilution of HRP-conjugated anti-human IgG (Sigma) for 1 hour at room temperature. Plates were washed and developed using TMB substrate (Sigma), stopped using sulphuric acid and read at 450nm. Endpoint titres were calculated as the reciprocal serum dilution giving signal 2x background using a fitted curve (4 parameter log regression).

### ACE2-RBD inhibition ELISA

An ELISA was performed to measure the ability of plasma antibodies to block interaction between recombinant human ACE2 and RBD proteins. 96-well Maxisorp plates (Thermo Fisher) were coated overnight at 4°C with 8 μg/ml of recombinant RBD protein in carbonate-bicarbonate coating buffer (Sigma). After blocking with PBS containing 1% BSA, duplicate wells of serially diluted plasma (1:25 to 1:1600) were added and incubated for 1 hour at room temperature. Plates were then incubated with 1.5 μg/ml of biotinylated recombinant ACE2 protein for 1 hour at room temperature followed by incubation with HRP-conjugated streptavidin (Thermo Fisher Scientific) for 1 hour at room temperature. Plates were developed with TMB substrate (Sigma), stopped with 0.15 M sulphuric acid and read at 450 nm.

### Microneutralisation Assay

SARS-CoV-2 isolate CoV/Australia/VIC01/2020^45^ was passaged in Vero cells and stored at −8°C. Plasma was heat-inactivated at 56°C for 30 min. Plasma was serially-diluted 1:20 to 1:10240 before addition of 100 TCID_50_ of SARS-CoV-2 in MEM/0.5% BSA and incubation at room temperature for 1 hour. Residual virus infectivity in the plasma/virus mixtures was assessed in quadruplicate wells of Vero cells incubated in serum-free media containing 1 μg/ml TPCK trypsin at 37°C/5% CO2; viral cytopathic effect was read on day 5. The neutralising antibody titre is calculated using the Reed/Muench method as previously described^21,22^.

### Flow cytometric detection of SARS-CoV-2 reactive B cells

Probes for delineating SARS-CoV-2 S-specific B cells within cryopreserved human PBMC were generated by sequential addition of streptavidin-PE (Thermofisher) to trimeric S protein biotinylated using recombinant Bir-A (Avidity). Biotinylated SARS-CoV RBD was similarly conjugated to streptavidin-BV421 (BD). SARS-CoV-2 RBD protein was directly labelled to APC using an APC Conjugation Lightning-link kit (Abcam). Cells were stained with Aqua viability dye (Thermofisher). Monoclonal antibodies for surface staining included: CD19-ECD (J3-119) (Beckman Coulter), CD20 Alexa700 (2H7), IgM-BUV395 (G20-127), CD21-BUV737 (B-ly4), IgD-Cy7PE (IA6-2), IgG-BV786 (G18-145) (BD), CD14-BV510 (M5E2), CD3-BV510 (OKT3), CD8a-BV510 (RPA-T8), CD16-BV510 (3G8), CD10-BV510 (HI10a), CD27-BV605 (O323) (Biolegend)._Cells were washed, fixed with 1% formaldehyde (Polysciences) and acquired on a BD LSR Fortessa or BD Aria II.

### Flow cytometric detection of antigen-specific cTFH and CD4 T cells

Cryopreserved human PBMC were thawed and rested for four hours at 37°C. Cells were cultured in 96-well plates at 1×10^6^ cells/well and stimulated for 20 hours with 5μg/mL of protein (BSA, SARS-CoV-2 S, SARS-CoV-2 RBD, HKU1 S). Selected donors were also stimulated with SEB (5μg/mL) as a positive control. Following stimulation, cells were washed, stained with Live/dead Blue viability dye (ThermoFisher), and a cocktail of monoclonal antibodies: CD27 BUV737 (L128), CCR7 Alexa700 (150503), CD45RA PeCy7 (HI100), CD20 BUV805 (2H7), CD14 BUV395 (MOP9) (BD Biosciences), CD3 BV510 (SK7), CD4 BV605 (RPA-T4), CD8 BV650 (RPA-T8), CD25 APC (BC96), OX-40 PerCP-Cy5.5 (ACT35), PD-1 BV421 (EH12.2H7), CCR6 BV785 (G034E3), CXCR3 Pe-Dazzle594 (G02H57) (Biolegend), and CXCR5 PE (MU5UBEE, ThermoFisher). Cells were washed, fixed with 1% formaldehyde and acquired on a BD LSR Fortessa using BD FACS Diva.

### Statistical Analyses

Grouped data are generally presented as median +/- IQR, with groups compared by Wilcoxon or Mann-Whitney U tests using Prism 8.0 (Graphpad). Pairwise correlations were assessed using Spearmans tests in Prism 8.0 (Graphpad). A multiple linear regression was used to determine which factors can be used to predict the values of microneutralization across 41 patients. A backward model selection method was used by first fitting a model with all variables of interest, and gradually removing one variable with the least significant p value. For each step, the nested F test was used to compare the models (ie: if the removed variable contributed significantly to the fit). This procedure was stopped when a model with only significant predictors was obtained. Confirmatory forward selection methods were also employed, arriving at the same final model. This analysis was performed using the *lm* function in *R* (v3.6.3).

Spearman correlation matrix was calculated using the *rcorr* function in *R* (v3.6.3). Principal component analysis was performed in *R* (v3.6.3) using the *princomp* function by first scaling the variables to have a unit variance. Throughout the manuscript, significance was defined as p<0.05.

## Data Availability

All data is available from the authors upon reasonable request

## Authors’ contributions

JAJ, HXT, WSL, SJK and AKW designed the study and experiments; JAJ, HXT, WSL, AR, HGK, KW, RE, HEK, CJB, FLM, NAG, PP, MD, NES and AKW performed experiments; WHT, NAG, DIG, KS contributed unique reagents; JAJ, HXT, WSL, AR, KS, MPD, SJK and AKW analysed the experimental data; JAJ, HXT, WSL, AR, MPD, SJK and AKW wrote the manuscript. All authors reviewed the manuscript.

## Acknowledgements

We thank the generous participation of the trial subjects for providing samples. The SARS-CoV-2 RBD expression plasmids were kindly provided by Florian Krammer, Mt Sinai School of Medicine, NY, USA. The human and mouse ACE2 expressionplasmids were kindly provided by Merlin Thomas, Monash University, Australia. We acknowledge the Melbourne Cytometry Platform (Melbourne Brain Centre node) for provision of flow cytometry services. We thank the Melbourne Mass Spectrometry and Proteomics Facility of The Bio21 Molecular Science and Biotechnology Institute at The University of Melbourne for the support of mass spectrometry analysis. This study was supported by the ARC Centre of Excellence in Convergent Bio-Nano Science and Technology (SJK), an NHMRC program grant APP1149990 (SJK), NHMRC project grant GNT1162760 (AKW), and the Jack Ma Foundation (DIG, NAG). WHT is a Howard Hughes Medical Institute-Wellcome Trust International Research Scholar (208693/Z/17/Z). JAJ, DIG, MPD, WHT, SJK and AKW are supported by NHMRC fellowships.

## SUPPLEMENTARY FIGURES

**Figure S1.**
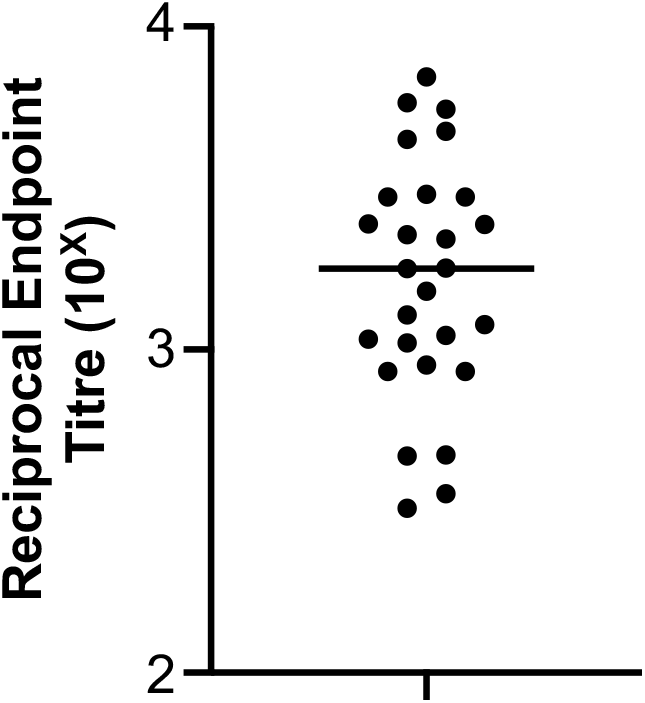
Pre-screen of HCoV-HKU1 serum endpoint titres among a cohort of healthy subjects (N=27) bled prior to the SARS-CoV-2 pandemic.

**Figure S2.**
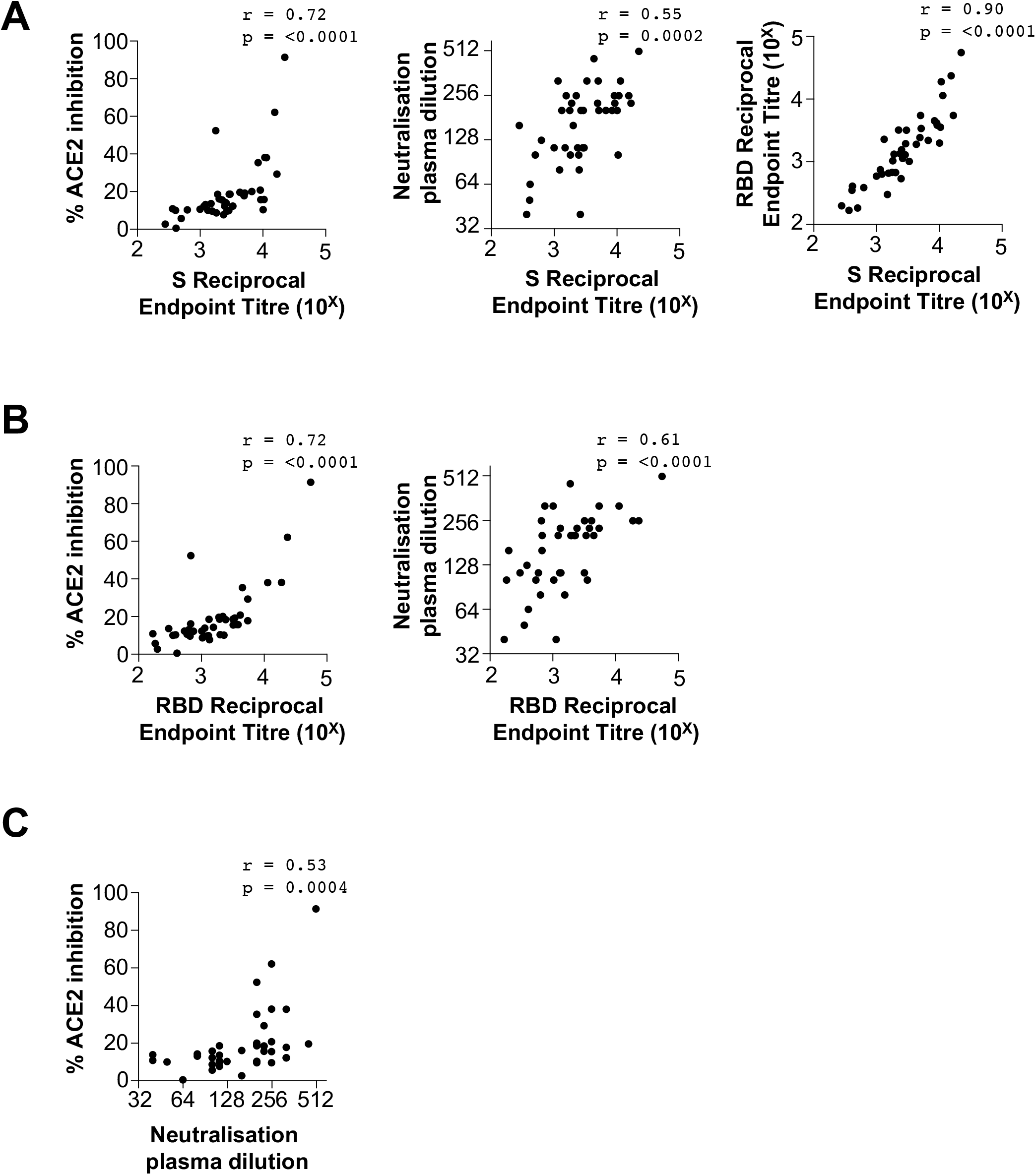
Correlations between antibody binding titres, ACE2/RBD binding inhibition and neutralisation activity in plasma from subjects recovered from SARSCoV-2 infection. (**A**) Correlation between endpoint titres of S-specific plasma antibody and the extent of ACE2/RBD binding inhibition, plasma neutralisation titres or RBD-specific plasma antibody. (**B**) Correlation between endpoint titres of RBD-specific plasma antibody and the extent of ACE2/RBD binding inhibition or plasma neutralisation titres. **(C)** Correlation between plasma ACE2/RBD binding inhibition and neutralisation activity.

**Figure S3.**
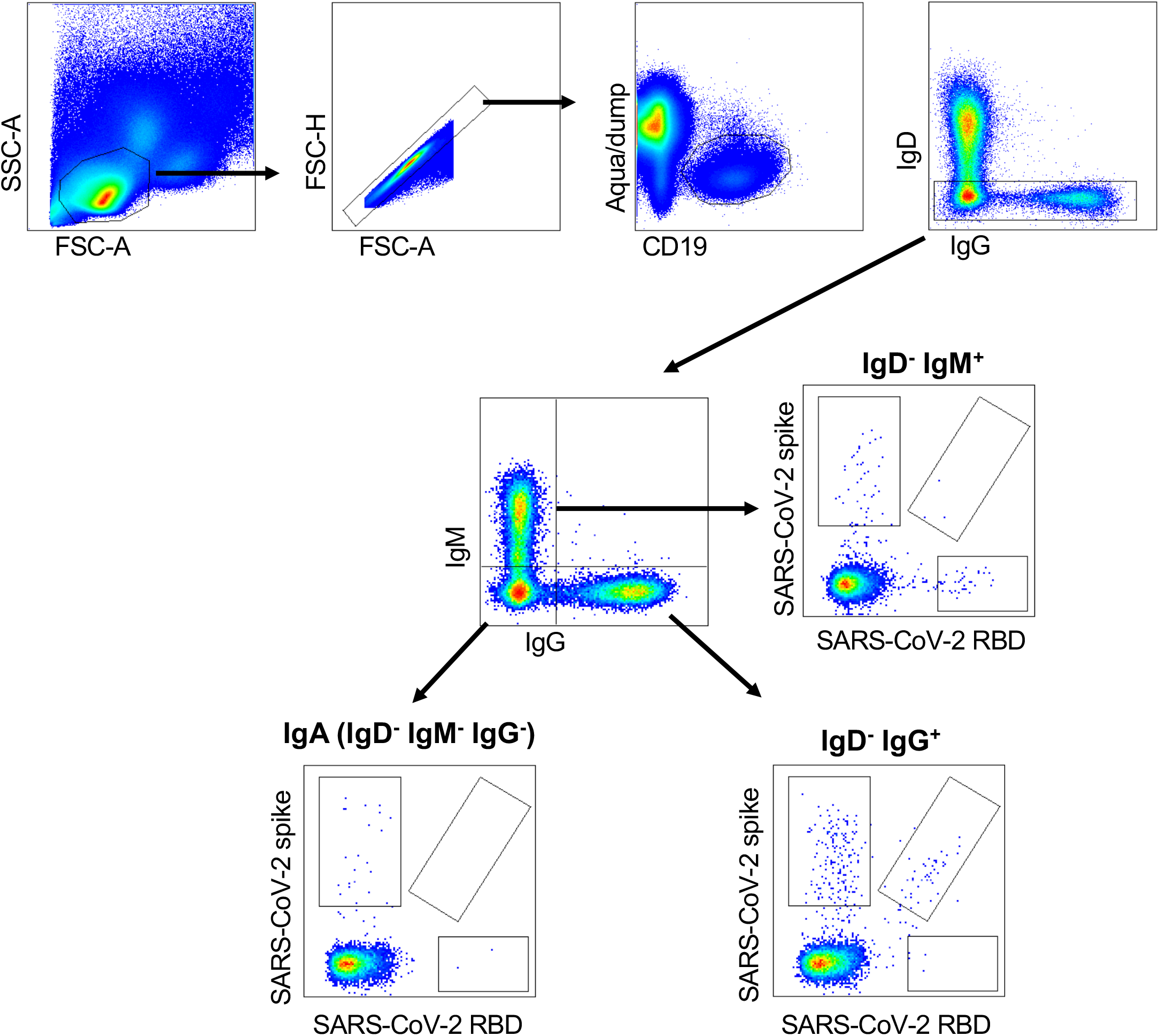
Gating strategy for resolving antigen-specific B cells and surface isotypes. Lymphocytes were identified by FSC-A vs SSC-A gating, followed by doublet exclusion (FSC-A vs FSC-H), and gating on live CD19^+^ B cells. Class-switched B cells were identified as IgD^−^, and surface istoype resolved by staining for IgM or IgG, with the double negative population (IgM^−^/IgG^−^) previously established as predominantly IgA. Binding to SARS-CoV-2 spike (S) and/or SARS-CoV-2 RBD probes was assessed for each population.

**Figure S4.**
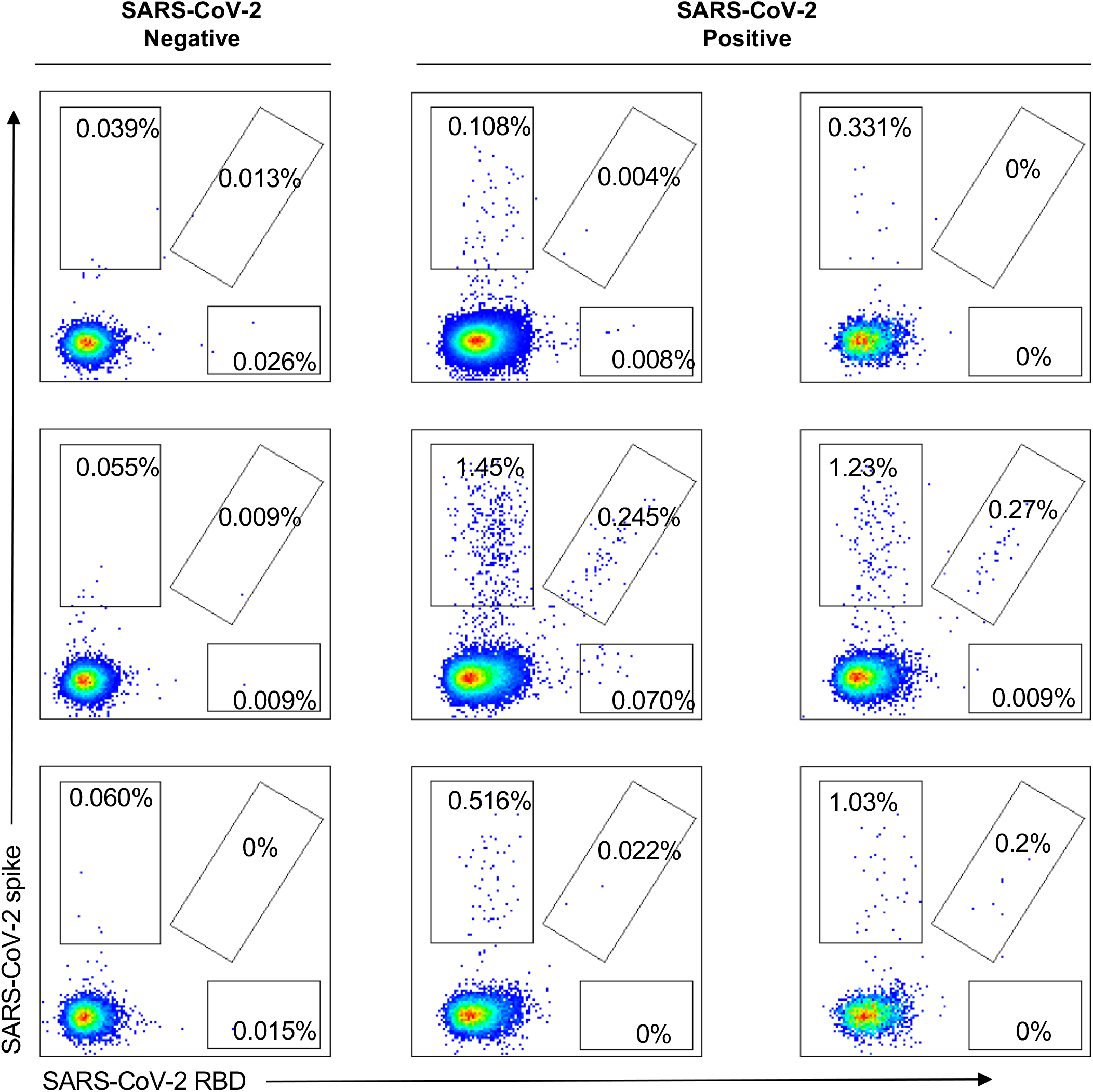
Representative staining of S- and RBD-specific IgD^−^IgG^+^ B cells. 3 uninfected subjects (left panels) and 6 subjects after recovery from SARS-CoV-2 infection (middle and right panels). CD19^+^IgD^−^IgG^+^ B cells cells were identified using gating strategy shown in Figure S5. Binding to SARS-CoV-2 spike (S) and/or SARS-CoV-2 RBD probes was assessed.

**Figure S5.**
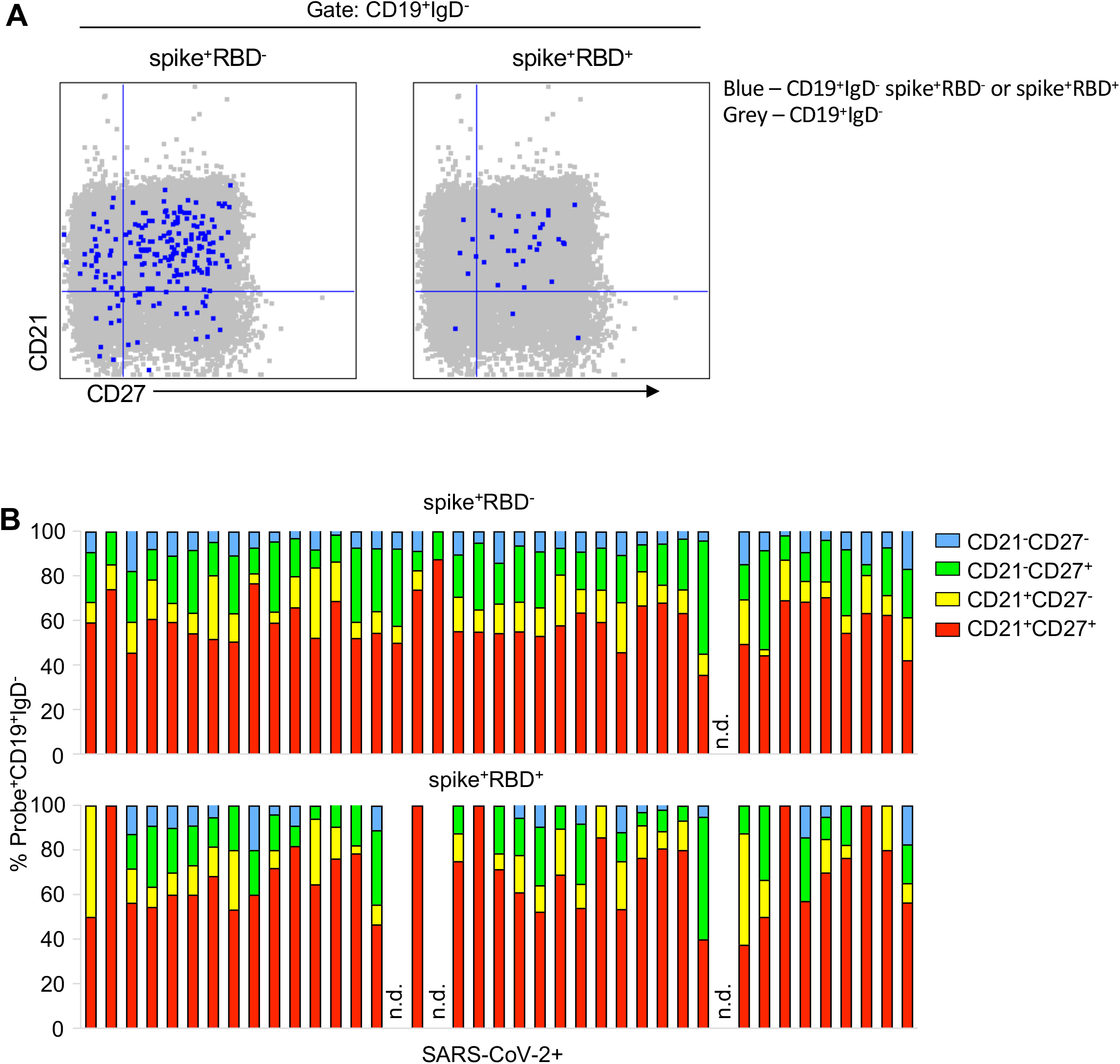
Memory B cell phenotypes in subjects after SARS-CoV-2 infection. **(A)** Representative memory B cell phenotypes identified by CD21 and CD27 co-stain of probe^+^CD19^+^IgD^−^ cells (blue) overlaid on CD19^+^IgD^−^ cells (grey) and **(B)** the corresponding frequencies of the four populations in subjects previously infected with SARS-CoV-2 (Resting memory – CD21^+^CD27^+^; activated memory – CD21^−^CD27^+^; naïve/CD27^lo^ memory – CD21^+^CD27^−^; atypical B cells – CD21^−^CD27^−^); n.d – not detected due to absent probe^+^ cells.

**Figure S6.**
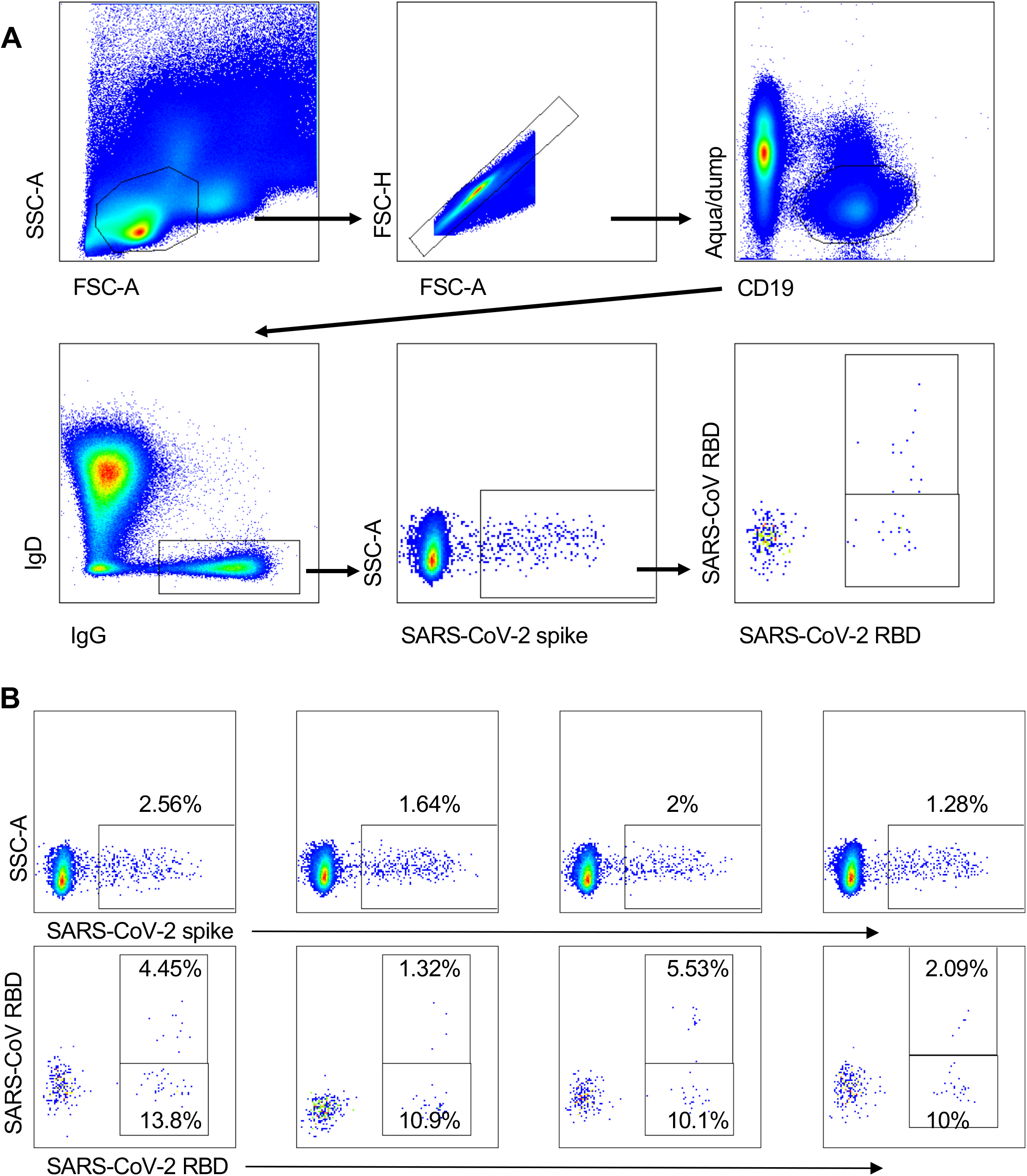
Gating strategy for resolving spike^+^ CD19^+^IgD^−^IgG^+^ B cells specific for SARS-CoV-2 and SARS-CoV RBD. (**A**) Lymphocytes were identified by FSC-A vs SSC-A gating, followed by doublet exclusion (FSC-A vs FSC-H), and gating on live CD19^+^ B cells. IgD^−^IgG^+^ B cells were gated and assessed for binding to SARS-CoV-2 spike. Cross-reactive specificities versus those unique to SARS-CoV-2 were discriminated by co-staining with SARS-CoV-2 and SARS-CoV RBD probes. (**B**) Representative staining shown for 4 subjects with prior SARS-CoV-2 infection.

**Figure S7.**
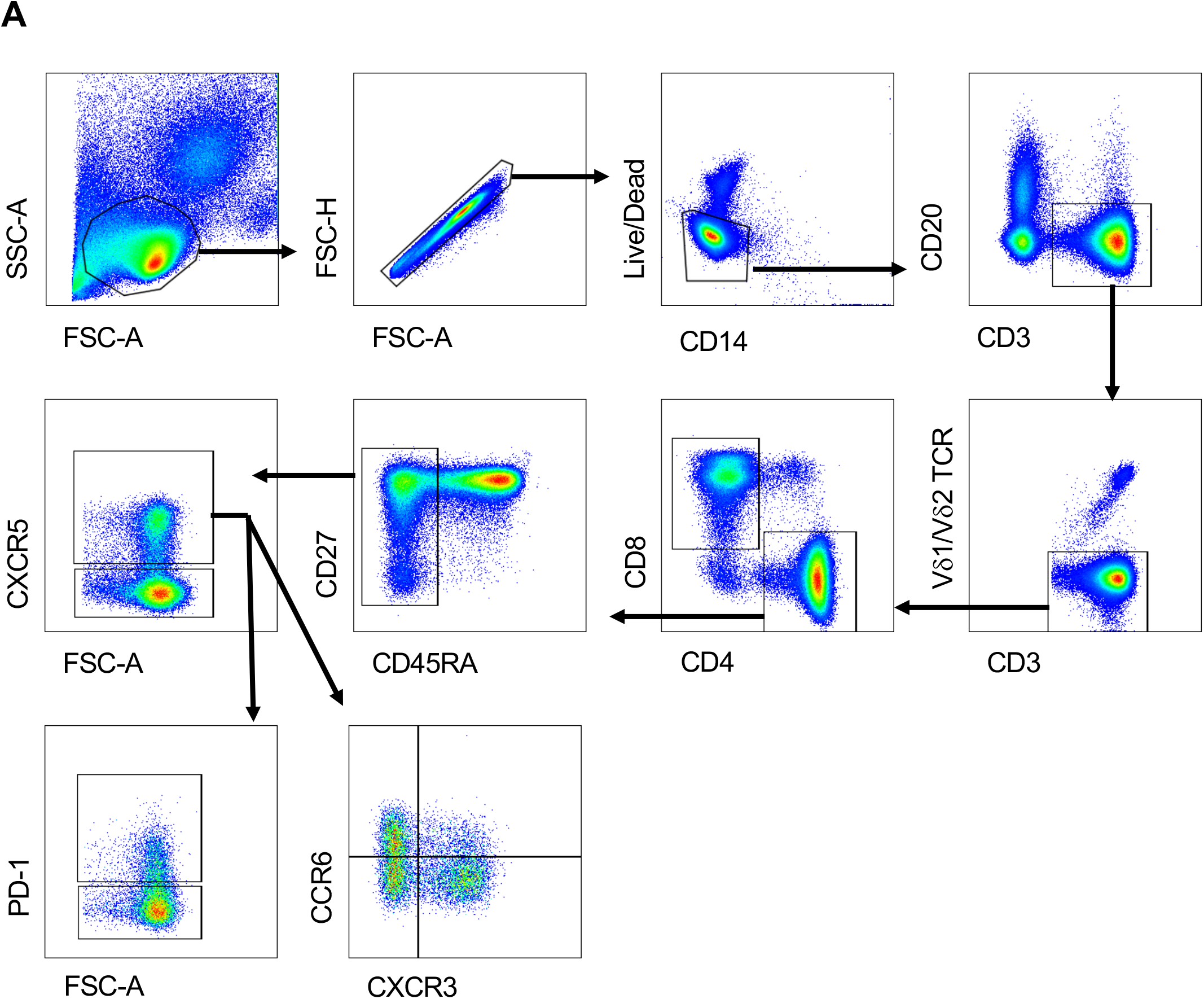
Gating strategy for cTFH and memory CD4^+^ T cell subsets. Lymphocytes were identified by FSC-A vs SSC-A gating, followed by doublet exclusion (FSC-A vs FSC-H gate), and exclusion of dead or CD14^+^ cells. T cells were identified as CD3^+^CD20^−^. Following exclusion of gamma delta T cells by Vd1/Vd2 TCR staining, CD4^+^CD8^−^ T cells were identified. Memory CD4^+^ T cells were defined as CD45RA^−^CXCR5^−^, while cTfh cells were defined as CD45RA^−^CXCR5^+^. cTFH cells were further characterized by PD-1 and CCR6/CXCR3 expression.

**Figure S8.**
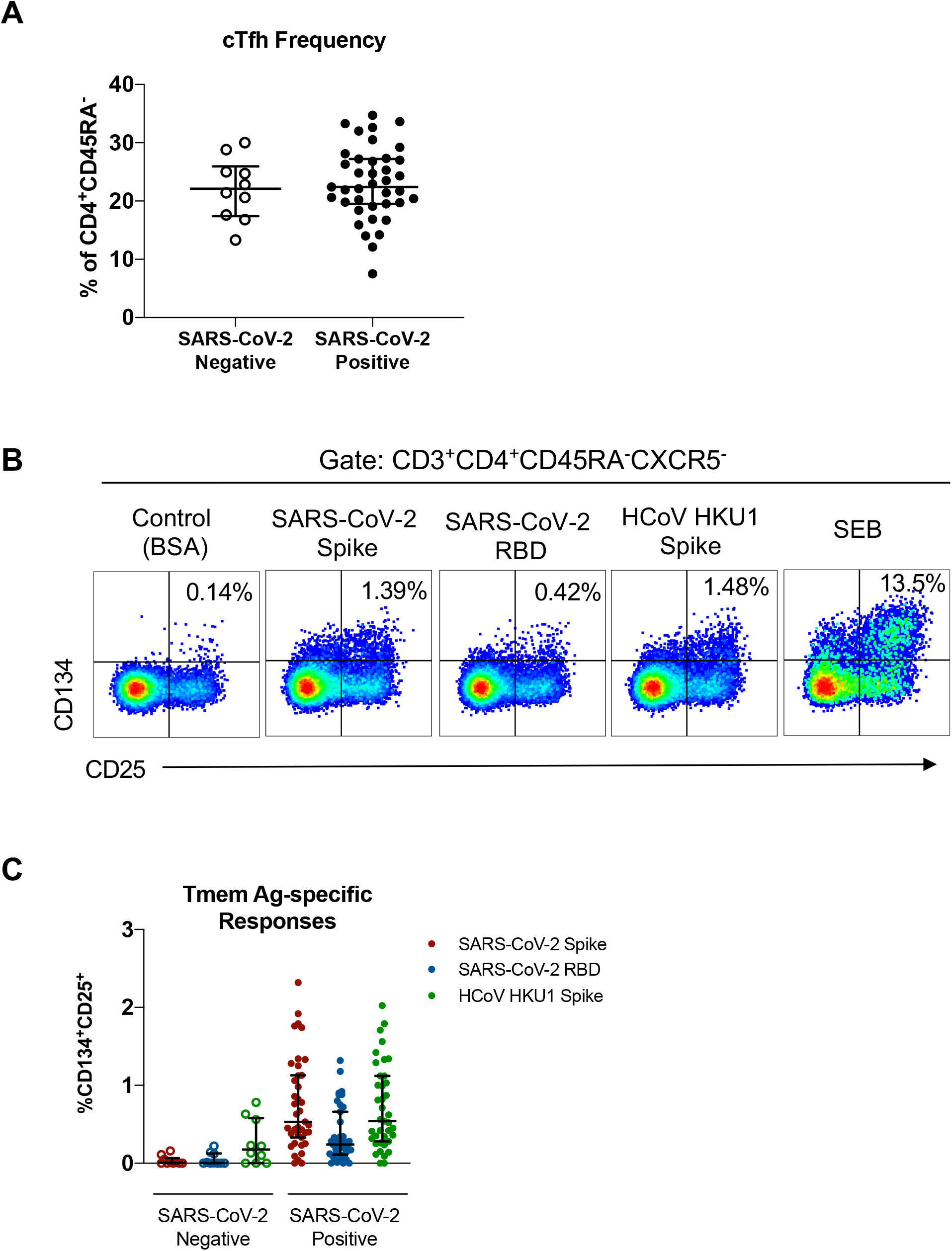
CD4 T cell populations in SARS-CoV-2 positive and negative donors. (**A**) cTFH frequency (as a proportion of CD4^+^CD45RA^−^ cells) among SARS-CoV-2 negative (n=10) and SARS-CoV-2 positive (n=39) donors. (**B**) Representative staining of CD134 and CD25 expression following protein or SEB stimulation among CD4 Tmem (CD4^+^CD45RA^−^CXCR5^−^) cells. (**C**) Background-subtracted frequencies of SARS-CoV-2 Spike, SARS-CoV-2 RBD or HCoV HKU1-specific CD4 Tmem cells among SARS-CoV-2 negative (n=10) or SARS-CoV-2 positive (n=39) donors.

**Figure S9.**
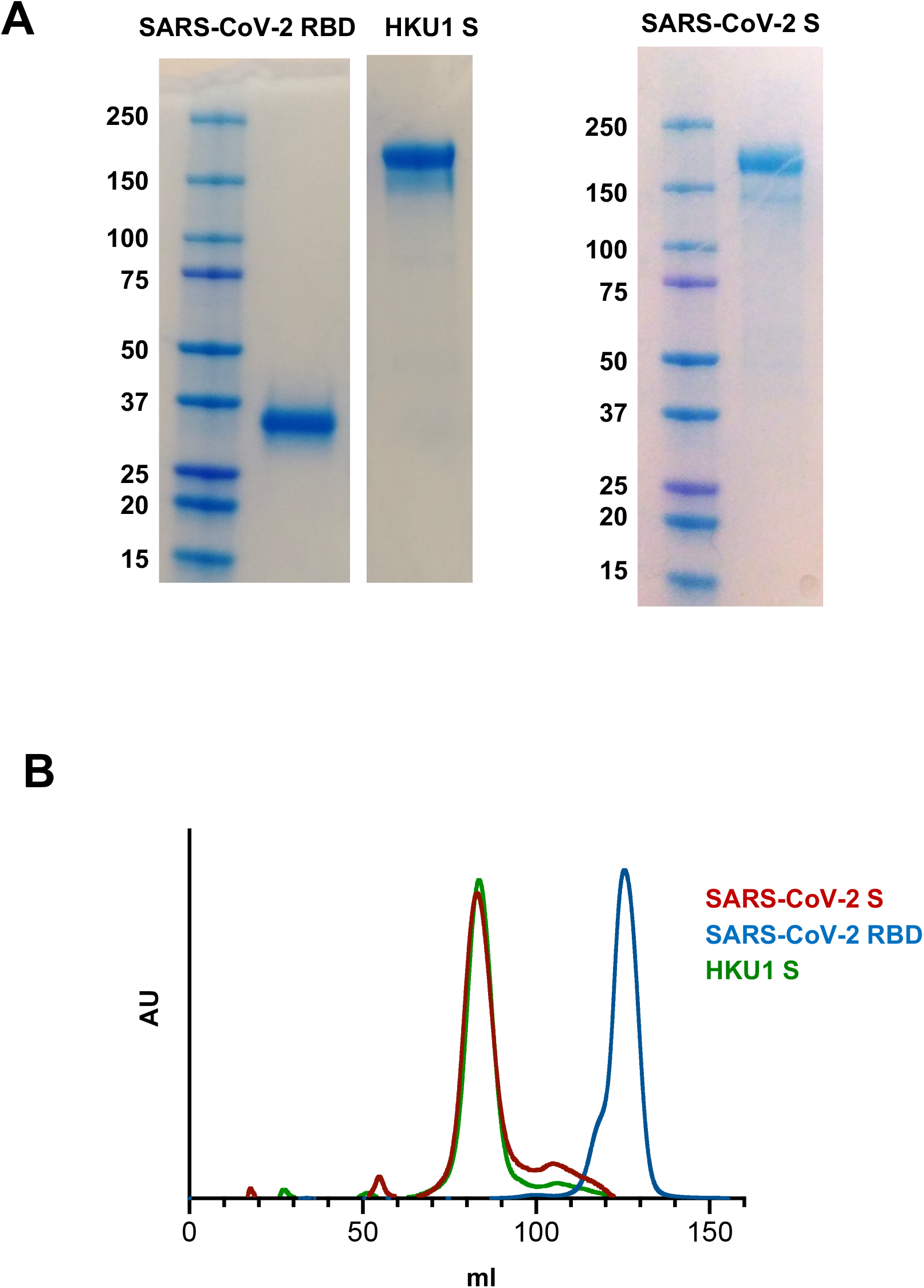
Purification of SARS-CoV-2 S, SARS-CoV-2 RBD and HCoV HKU1 S proteins. S proteins for SARS-CoV-2 and HCoV-HKU1, and the RBD domain of SARS-CoV-2 were expressed in Expi293 cells and purified using Ni-NTA and size exclusion chromatography. (**A**) SDS-PAGE of purified proteins stained with Coomassie. (**B**) Gel filtration profiles of recombinant coronaviral proteins run on a Superose 6 16/70 column.

**Figure S10.**
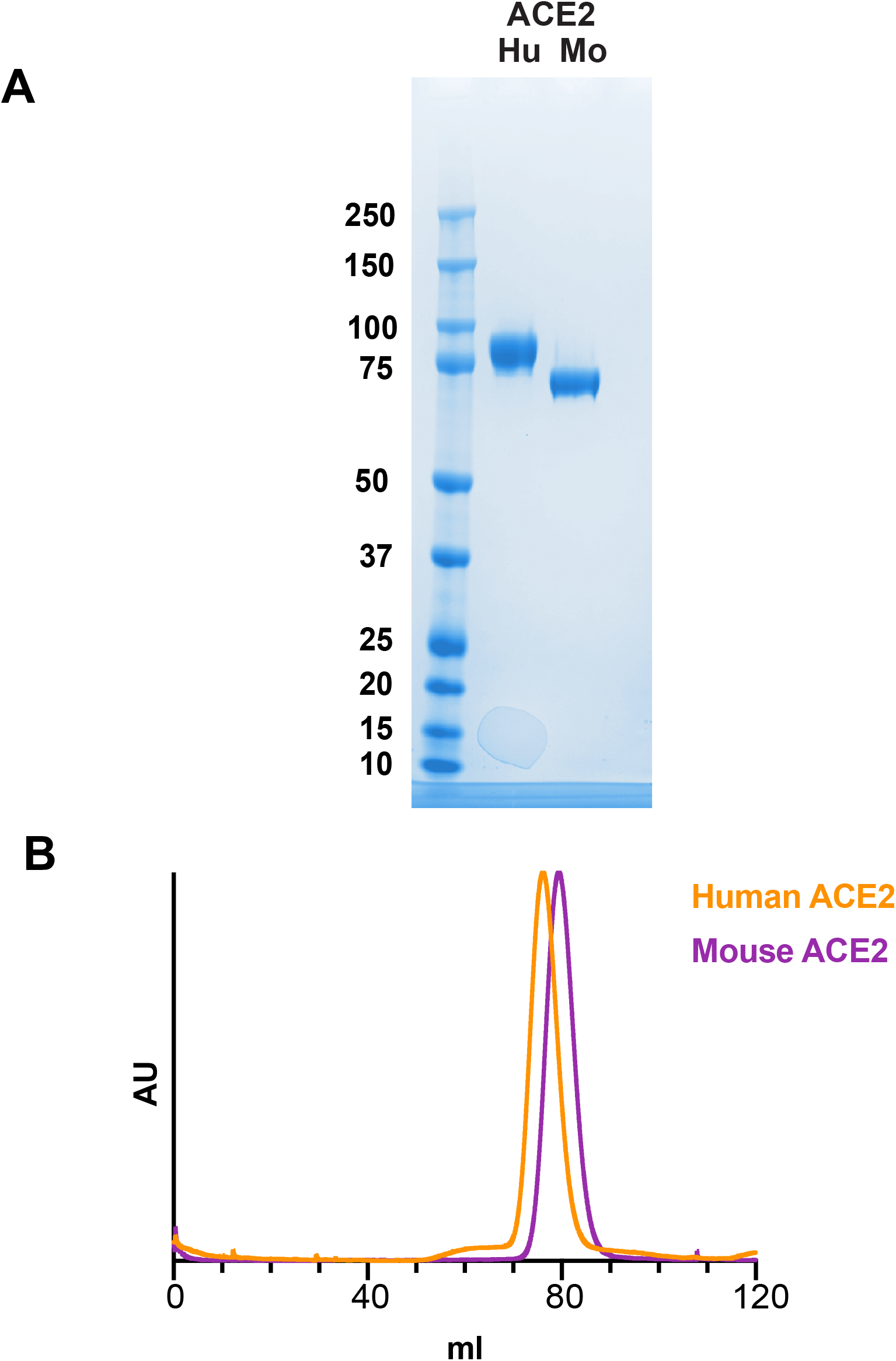
Purification of human and mouse ACE2 proteins. Proteins were expressed in Expi293 cells and purified using Ni-NTA and size exclusion chromatography. (**A**) SDS-PAGE of purified proteins stained with Coomassie. (**B**) Gel filtration profiles of recombinant proteins run on a Hiload 16/60 Superdex 200 Prep Grade column.

**Figure S11.**
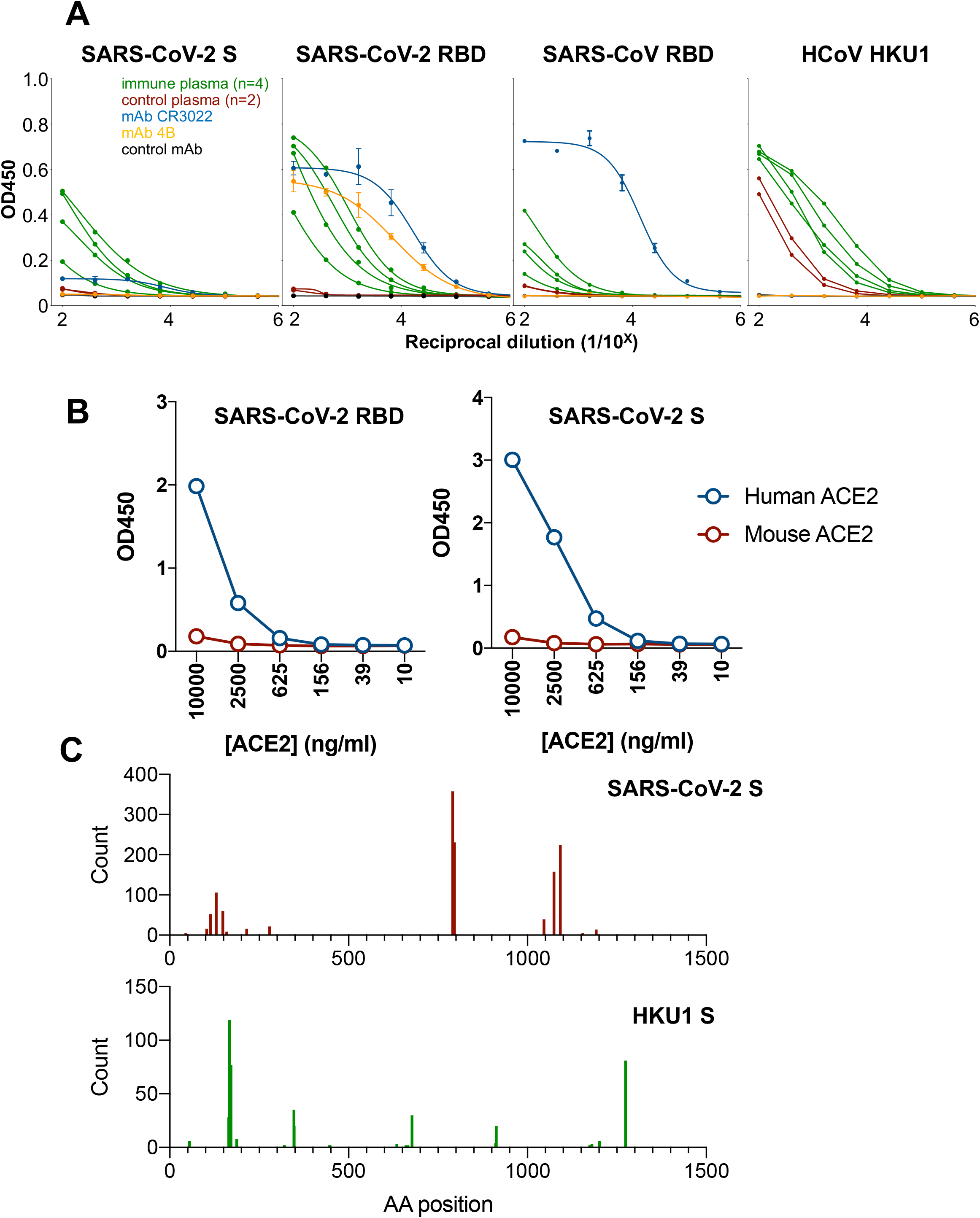
Antigenicity and glycosylation profile of coronavirus proteins. (**A**) Plates were coated overnight with 2ug/ml of SARS-CoV-2 S, SARS-CoV-2 RBD, SARS-CoV RBD or HCoV-HKU1. Binding to control plasma (red), plasma from recovered SARS-CoV-2 infected individuals (green) or monoclonal antibodies CR3022 (blue), 4B (orange) or control CR8071 (black) was assessed by ELISA. (**B**) Binding of SARS-CoV-2 S and SARS-CoV-2 RBD to recombinant human but not mouse ACE2 was established by ELISA. (**C**) Glycan occupancy of SARS-CoV-2 S and HCoV-HKU1 S was assessed using mass spectrometry.

**Table S1.** Characteristics of 41 subjects recovered from COVID-19.

| Characteristic | Median | IQR |
| --- | --- | --- |
| Age, median - years | 59 | (54-65) |
| Male sex – no. (%) | 24 (59%) |  |
| Time since symptom onset - days | 36 | (32-40) |
| Time since +ve nasal swab SAR-CoV-2 PCR* - days | 32 | (28-35) |
| Illness severity** |  |  |
| Mild – no. (%) | 26 (63.4%*) |  |
| Moderate – no. (%) | 10 (24.4%) |  |
| Severe – no. (%) | 5 (12.2%) |  |

\* 3 Subjects had a compatible illness and history of exposure but did not have a positive nasal swab
\*\* Illness severity was classified as:
Mild: prominent upper respiratory tract symptoms and not hospitalised.
Moderate: prominent lower respiratory tract symptoms and not hospitalised.
Severe: prominent lower respiratory tract symptoms and requiring hospital care.

**Table S2.**
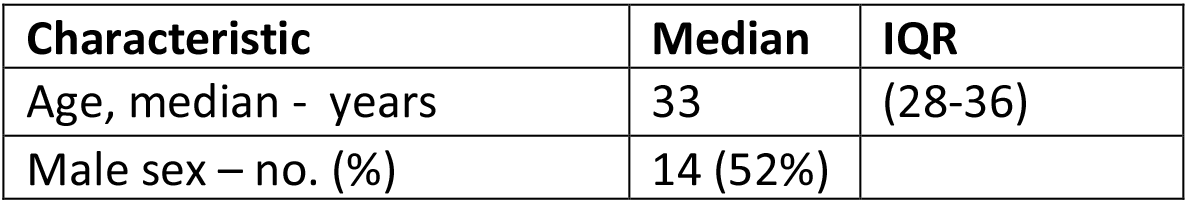
Age and sex of 27 healthy control subjects without COVID-19 symptoms.

| Characteristic | Median | IQR |
| --- | --- | --- |
| Age, median - years | 33 | (28-36) |
| Male sex – no. (%) | 14 (52%) |  |

## References

1. Li, F. Structure, Function, and Evolution of Coronavirus Spike Proteins. Annu Rev Virol 3, 237–261 (2016).

2. Hoffmann, M., et al. SARS-CoV-2 Cell Entry Depends on ACE2 and TMPRSS2 and Is Blocked by a Clinically Proven Protease Inhibitor. Cell 181, 271–280.e278 (2020).

3. Yan, R., et al. Structural basis for the recognition of SARS-CoV-2 by full-length human ACE2. Science (New York, N.Y.) 367, 1444–1448 (2020).

4. Lan, J., et al. Structure of the SARS-CoV-2 spike receptor-binding domain bound to the ACE2 receptor. Nature (2020).

5. Jiang, S., Hillyer, C. & Du, L. Neutralizing Antibodies against SARS-CoV-2 and Other Human Coronaviruses. Trends in immunology 41, 355–359 (2020).

6. Tian, X., et al. Potent binding of 2019 novel coronavirus spike protein by a SARS coronavirus-specific human monoclonal antibody. Emerg Microbes Infect 9, 382–385 (2020).

7. Yuan, M., et al. A highly conserved cryptic epitope in the receptor binding domains of SARS-CoV-2 and SARS-CoV. Science (New York, N.Y.) 368, 630–633 (2020).

8. Pinto, D., et al. Structural and functional analysis of a potent sarbecovirus neutralizing antibody. *bioRxiv*, 2020.2004.2007.023903 (2020).

9. Ju, B., et al. Potent human neutralizing antibodies elicited by SARS-CoV-2 infection. *bioRxiv*, 2020.2003.2021.990770 (2020).

10. Chi, X., et al. A potent neutralizing human antibody reveals the N-terminal domain of the Spike protein of SARS-CoV-2 as a site of vulnerability. *bioRxiv*, 2020.2005.2008.083964 (2020).

11. Wang, L., et al. Importance of Neutralizing Monoclonal Antibodies Targeting Multiple Antigenic Sites on the Middle East Respiratory Syndrome Coronavirus Spike Glycoprotein To Avoid Neutralization Escape. Journal of virology 92(2018).

12. Widjaja, I., et al. Towards a solution to MERS: protective human monoclonal antibodies targeting different domains and functions of the MERS-coronavirus spike glycoprotein. Emerg Microbes Infect 8, 516–530 (2019).

13. Sui, J., et al. Potent neutralization of severe acute respiratory syndrome (SARS) coronavirus by a human mAb to S1 protein that blocks receptor association. Proceedings of the National Academy of Sciences of the United States of America 101, 2536–2541 (2004).

14. Lip, K.M., et al. Monoclonal antibodies targeting the HR2 domain and the region immediately upstream of the HR2 of the S protein neutralize in vitro infection of severe acute respiratory syndrome coronavirus. Journal of virology 80, 941–950 (2006).

15. Amanat, F. & Krammer, F. SARS-CoV-2 Vaccines: Status Report. Immunity 52, 583–589 (2020).

16. Thanh Le, T., et al. The COVID-19 vaccine development landscape. Nature reviews. Drug discovery 19, 305–306 (2020).

17. Gao, Q., et al. Rapid development of an inactivated vaccine candidate for SARS-CoV-2. Science (New York, N.Y.) (2020).

18. Lv, H., et al. Cross-reactive antibody response between SARS-CoV-2 and SARS-CoV infections. *bioRxiv*, 2020.2003.2015.993097 (2020).

19. Zhou, W., Wang, W., Wang, H., Lu, R. & Tan, W. First infection by all four non-severe acute respiratory syndrome human coronaviruses takes place during childhood. BMC infectious diseases 13, 433 (2013).

20. Okba, N.M.A., et al. Severe Acute Respiratory Syndrome Coronavirus 2-Specific Antibody Responses in Coronavirus Disease 2019 Patients. Emerg Infect Dis 26(2020).

21. Subbarao, K., et al. Prior infection and passive transfer of neutralizing antibody prevent replication of severe acute respiratory syndrome coronavirus in the respiratory tract of mice. Journal of virology 78, 3572–3577 (2004).

22. Houser, K.V., et al. Prophylaxis With a Middle East Respiratory Syndrome Coronavirus (MERS-CoV)-Specific Human Monoclonal Antibody Protects Rabbits From MERS-CoV Infection. The Journal of infectious diseases 213, 1557–1561 (2016).

23. Koutsakos, M., et al. Circulating T(FH) cells, serological memory, and tissue compartmentalization shape human influenza-specific B cell immunity. Science translational medicine 10(2018).

24. Lau, D., et al. Low CD21 expression defines a population of recent germinal center graduates primed for plasma cell differentiation. Science immunology 2(2017).

25. Ou, X., et al. Characterization of spike glycoprotein of SARS-CoV-2 on virus entry and its immune cross-reactivity with SARS-CoV. Nature communications 11, 1620 (2020).

26. Crotty, S. T Follicular Helper Cell Biology: A Decade of Discovery and Diseases. Immunity 50, 1132–1148 (2019).

27. Brenna, E., et al. CD4(+) T Follicular Helper Cells in Human Tonsils and Blood Are Clonally Convergent but Divergent from Non-Tfh CD4(+) Cells. Cell reports 30, 137–152.e135 (2020).

28. Morita, R., et al. Human blood CXCR5(+)CD4(+) T cells are counterparts of T follicular cells and contain specific subsets that differentially support antibody secretion. Immunity 34, 108–121 (2011).

29. Dan, J.M., et al. A Cytokine-Independent Approach To Identify Antigen-Specific Human Germinal Center T Follicular Helper Cells and Rare Antigen-Specific CD4+ T Cells in Blood. Journal of immunology (Baltimore, Md.: 1950) 197, 983–993 (2016).

30. Bentebibel, S.E., et al. Induction of ICOS+CXCR3+CXCR5+ TH cells correlates with antibody responses to influenza vaccination. Science translational medicine 5, 176ra132 (2013).

31. Koutsakos, M., Nguyen, T.H.O. & Kedzierska, K. With a Little Help from T Follicular Helper Friends: Humoral Immunity to Influenza Vaccination. Journal of immunology (Baltimore, Md.: 1950) 202, 360–367 (2019).

32. Brouwer, P., et al. Potent neutralizing antibodies from COVID-19 patients define multiple targets of vulnerability. *bioRxiv*, 2020.2005.2012.088716 (2020).

33. Tan, H.X., et al. Subdominance and poor intrinsic immunogenicity limit humoral immunity targeting influenza HA stem. The Journal of clinical investigation 129, 850–862 (2019).

34. Farooq, F., et al. Circulating follicular T helper cells and cytokine profile in humans following vaccination with the rVSV-ZEBOV Ebola vaccine. Scientific reports 6, 27944 (2016).

35. Bentebibel, S.E., et al. ICOS(+)PD-1(+)CXCR3(+) T follicular helper cells contribute to the generation of high-avidity antibodies following influenza vaccination. Scientific reports 6, 26494 (2016).

36. Niessl, J., et al. Persistent expansion and Th1-like skewing of HIV-specific circulating T follicular helper cells during antiretroviral therapy. EBioMedicine 54, 102727 (2020).

37. Zhang, J., et al. Circulating CXCR3(+) Tfh cells positively correlate with neutralizing antibody responses in HCV-infected patients. Scientific reports 9, 10090 (2019).

38. Braun, J., et al. Presence of SARS-CoV-2 reactive T cells in COVID-19 patients and healthy donors. *medRxiv*, 2020.2004.2017.20061440 (2020).

39. Wrapp, D., et al. Cryo-EM structure of the 2019-nCoV spike in the prefusion conformation. Science (New York, N.Y.) 367, 1260–1263 (2020).

40. Amanat, F., et al. A serological assay to detect SARS-CoV-2 seroconversion in humans. Nature medicine (2020).

41. Lopez, E., et al. Low pH Exposure During Immunoglobulin G Purification Methods Results in Aggregates That Avidly Bind Fcγ Receptors: Implications for Measuring Fc Dependent Antibody Functions. Frontiers in immunology 10, 2415 (2019).

42. Hughes, C.S., et al. Single-pot, solid-phase-enhanced sample preparation for proteomics experiments. Nat Protoc 14, 68–85 (2019).

43. Bern, M., Kil, Y.J. & Becker, C. Byonic: advanced peptide and protein identification software. Curr Protoc Bioinformatics Chapter 13, Unit13.20 (2012).

44. Lee, L.Y., et al. Toward Automated N-Glycopeptide Identification in Glycoproteomics. Journal of proteome research 15, 3904–3915 (2016).

45. Caly, L., et al. Isolation and rapid sharing of the 2019 novel coronavirus (SARS-CoV-2) from the first patient diagnosed with COVID-19 in Australia. Med J Aust (2020).

